# Lineage B Genotype III of Dengue Virus Serotype 3 (DENV-3III_B) is responsible for Dengue Outbreak in Dire Dawa City, Ethiopia, 2023

**DOI:** 10.1101/2024.08.22.24312404

**Authors:** Abebe Asefa Negeri, Dawit Hailu Alemayehu, Saro Abdella Abrahim, Tsigereda Kifle Wolde, Gutema Bulti, Alemnesh Hailemariam Bedasso, Danile Tsega, Ebise Abosex, Eyilachew Zenebe Awule, Diana Rojas-Gallardo, Asefa Konde Korkiso, Kalkidan Melaku, Raffael Joseph, Abaysew Ayele, Mesfin Mengesha Tsegaye, Anne Piantadosi, Getachew Tollera, Alemseged Abdissa, Mesay Hailu Dangisso, Adane Mihret, Andargachew Mulu, Tesfaye Gelanew

## Abstract

**Background:** The eastern parts of Ethiopia, including Dire Dawa City, have faced annual dengue fever (DF) outbreaks since 2013, resulting in substantial healthcare and economic consequences. However, there remains a lack of comprehensive evidence regarding the specific dengue virus (DENV) serotypes and genotypes associated with those outbreaks.

**Methodology:** On December 14, 2023, the National Arbovirus Laboratory at the Ethiopian Public Health Institute received seventy serum samples from 70 patients suspected of DF during the outbreak in Dire Dawa City. Samples positive for DENV with adequate volume and CT values underwent sequencing of the CprM region of the DENV genome. The obtained sequences were typed using the Dengue Virus Typing Tool and underwent phylogenetic analysis to evaluate sequence diversity and relationships to DENV strains from other global DENV-endemic regions.

**Results:** Overall, 32 (45.7%) of the patients displayed one or more early warning signs indicative of severe dengue. Among the 13 patients who were hospitalized for 2 to 10 days, all except two had at least one symptom indicative of severe dengue. Out of 67 samples with adequate volume, 44 (65.6%) were positive for DENV RNA using RT-PCR, and 21 successfully underwent CprM sequencing. All of them belonged to DENV-3, genotype III, major lineage B (DENV-3III_B), but represented two different minor lineages (DENV-3III_B.2 and DENV-3III_B.3). Phylogenetic analysis demonstrated that both lineages were closely related to sequences from the Afar region of Ethiopia collected in 2023, indicating that the outbreaks were related and were due to multiple co-circulating lineages.

**Conclusions:** DENV-3III_B was identified as the cause of the 2023 DF outbreak in Dire Dawa City. The identification of two co-circulating lineages in this outbreak underscores the complex nature of DENV transmission in the African continent and highlights the need for increased viral genomic surveillance.

## 1. Introduction

Dengue fever (DF), caused by four genetically distinct serotypes of dengue virus (DENV-1 to DENV-4), has become a significant global health challenge. Over the past two decades, the incidence of dengue has increased tenfold, rising from 500,000 reported cases worldwide in 2000 to a staggering 5.2 million cases in 2019. In 2019, dengue outbreaks spread across 129 countries, marking an unprecedented peak. Although there was a slight decline in cases between 2020 and 2022 due to the COVID-19 pandemic and reduced reporting rates, 2023 witnessed a resurgence in dengue cases globally[1],[2],[3].

In areas where DF cases have been identified as endemic and/or epidemic, especially in tropical and subtropical countries, nonspecific febrile illnesses are common. Febrile illnesses can be caused by a variety of infectious agents such as malaria parasites, alphaviruses, and flaviviruses, complicating surveillance and response programs for outbreaks and endemic diseases[4]. In addition, the *Aede*s mosquito species, particularly *Ae. Aegypti* and *Ae. Albopictus* that transmit DENV, are also responsible for the transmission of other arboviruses (e.g., Zika virus, Yellow Fever virus, and Chikungunya virus), and are common in tropical and subtropical countries[5].

In Ethiopia, the first laboratory-confirmed outbreak of DF occurred in Deri Dawa City in 2013. Since then, Ethiopia has experienced nearly annual outbreaks of DF in multiple regions of the country (Figure 1). In the Afar Region, DF outbreaks occurred in 2019 in Gewane District[6]. In the Ethiopian Somalia region, the first outbreak was reported in Godey town between 2014 and 2015 followed by outbreaks that occurred in Kabridahar in 2017[7] and Warder Woreda in 2021[8]. These recurrent outbreaks have impacted an already fragmented health system[9] and the economy of these parts of the country.

**Figure 1.**
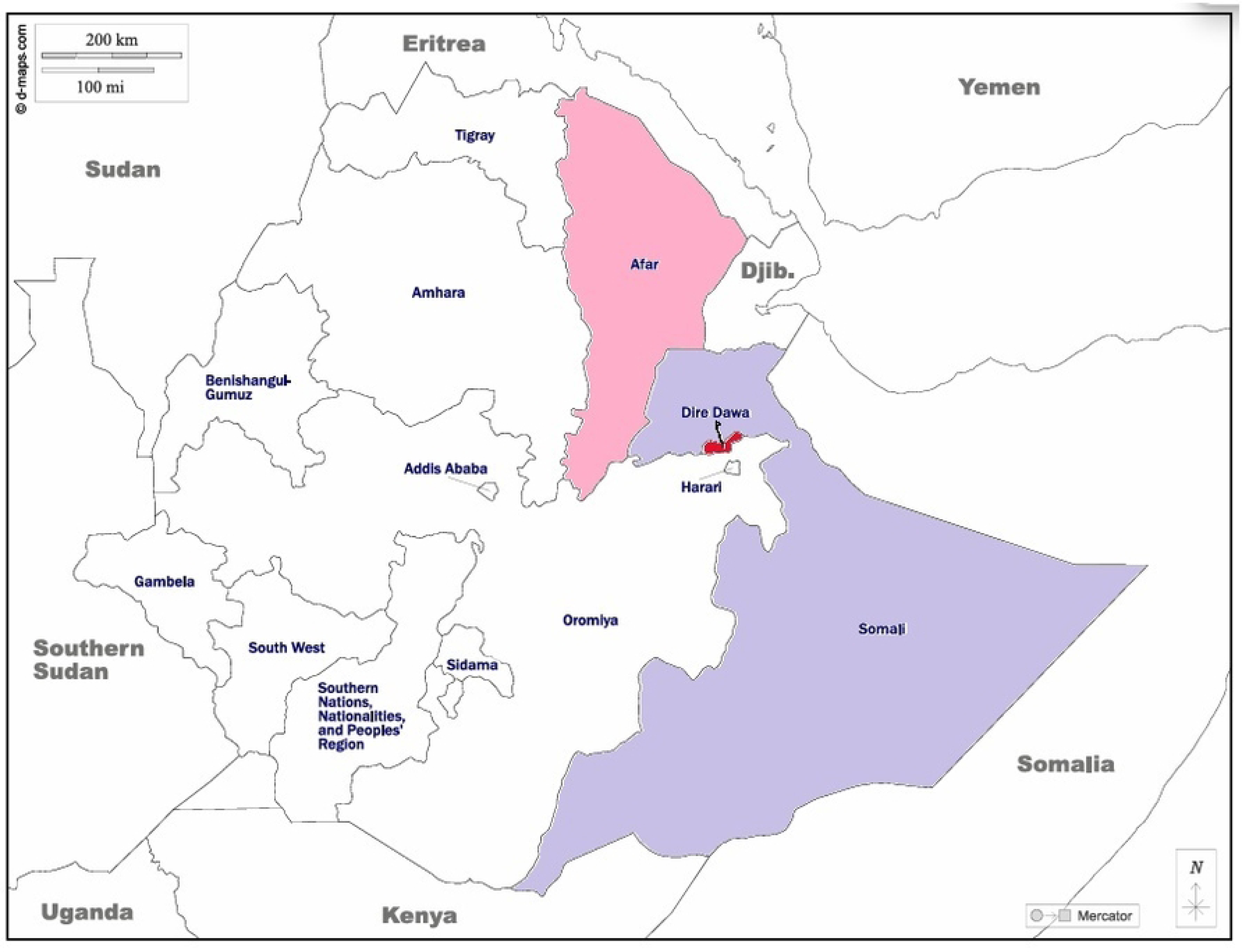
Map showing the Dire Dawa City Administration where the Dengue outbreak occurred, and study DENV strains originated. The shaded areas indicate the epicenters of dengue outbreaks in Ethiopia since 2013: Dire Dawa (red), Afar (red-violet), and Somalia (light blue)

In April 2023, an outbreak of DF was initially reported in the Afar region, the northeastern part of Ethiopia. The first affected districts were Logia and Mille. Since April 4, 2023, however, it spread to all seven districts and towns in the Afar region (Figure 1). As of June 26, 2023, a total of 6,133 suspected and confirmed cases were reported, resulting in nine associated deaths (with a case: fatality ratio of 0.5%)[10]. It was documented that 8 of the 10 epidemic samples examined by RT-PCT triplex [for Dengue, Zika, and Chikungunya] belonged to serotype DENV-3 [10].

As a suspected expansion of the outbreak in the Afar region, a DF outbreak was recorded in Dire Dawa City in December 2023 (Figure 1). However, there is no data regarding the DENV serotypes and genotypes associated with the outbreak. Whether the outbreak was caused by DENV-3 as in the Afar region remains unknown. In the present study, we present data on the serotypes, genotypes, and lineages of DENV strains responsible for the outbreak in Dire Dawa City during 2023 and discuss the epidemiological and clinical consequences.

## 2. Materials and Methods

### 2.1. Ethical considerations

As our study aimed to better understand DENV strains linked to DF outbreaks, we obtained an ethical approval waiver (protocol number: PO-036-24) from the All-African Leprosy Rehabilitation and Training Center/Armauer Hansen Research (ALERT/AHRI) Ethics Committee. Additionally, we obtained permission from the Dire Dawa City Health Bureau and health facilities’ authorities to investigate these outbreak samples. Since the samples used in this study were obtained from the dengue outbreak during the 2023 outbreak investigation and confirmation undertaken to respond to and control the public health emergencies, no informed consent was obtained during the sample collection.

### 2.2. Study Settings and Patient Data

On December 14, 2023, the Ethiopia Public Health Institute (EPH) received 70 serum samples from an outbreak of DF in Dire Dawa City Administration (Figure 1). These samples were collected from DF suspected cases and shipped to the National Arbovirus Laboratory of EPHI for laboratory confirmation of the causative agent as part of an outbreak response investigation (Figure 2). The reported clinical signs and symptoms for each suspected case were extracted from medical charts available at the health facilities they were admitted using a case-based surveillance request form. Mild symptoms include sudden onset of high fever, vomiting, nausea, rash, arthralgia (joint pain), headache, chills, myalgia (muscle pain), sore throat, dizziness, runny nose, pruritus (itchiness), adenosis, diarrhea and shivering while warning signs or indicative of severe dengue include: belly pain or tenderness, persistent vomiting (at least 3 times/24 hrs), bleeding from the nose or gum, restless or irritable, feeling weak, conjunctiva congestion and convulsions (Supplementary Table 1 and Table 2). The category of mild symptoms and warning signs indicative of severe dengue is based on the WHO severe dengue classification[11].

**Figure 2.**
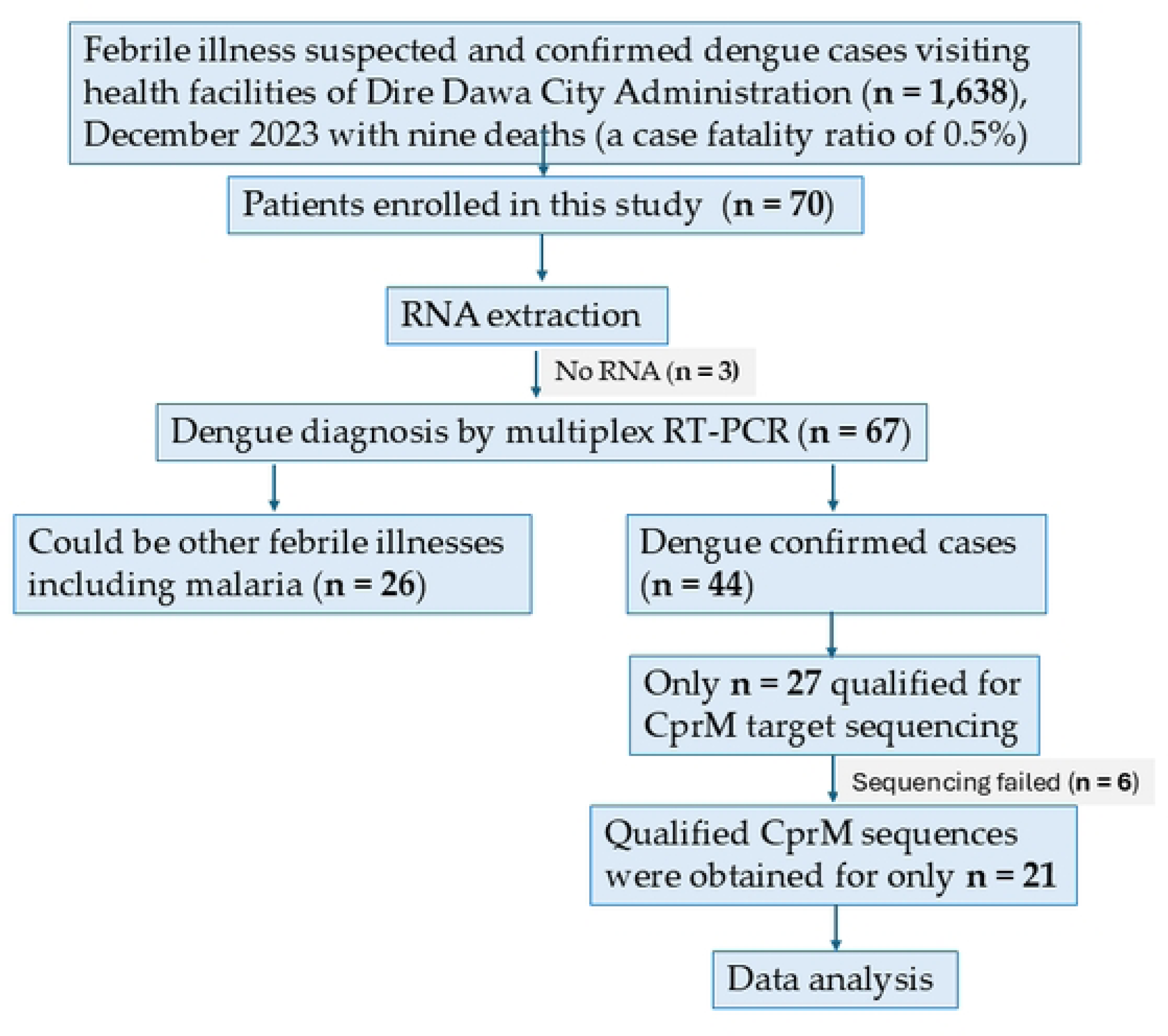
Flow chart showing enrollment of suspected Dengue and/ or other febrile illnesses, including malaria patients and molecular detection and characterization of strains isolated during the 2023 dengue outbreak in Dire Dawa, Ethiopia.

**Table 2.**
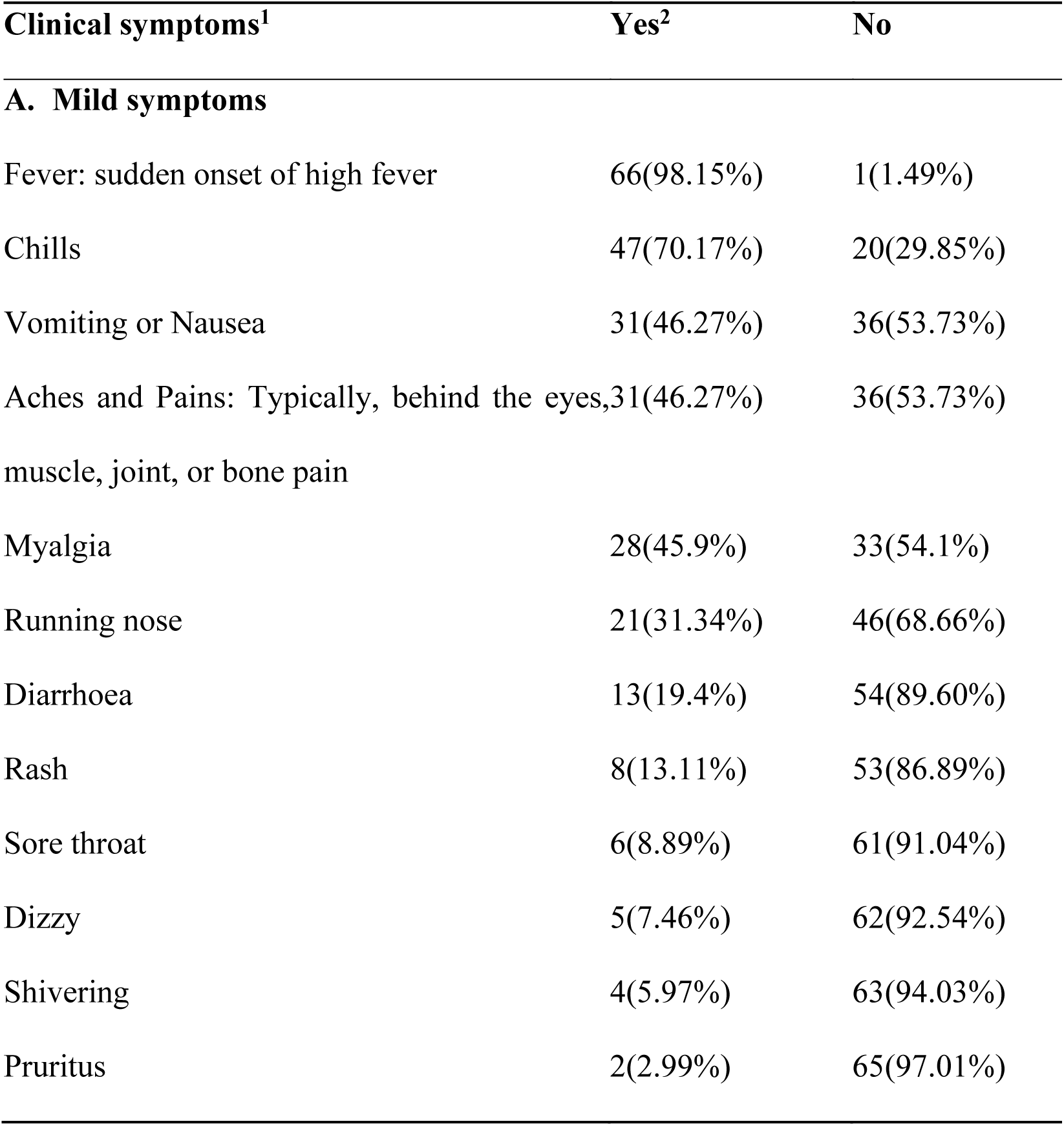
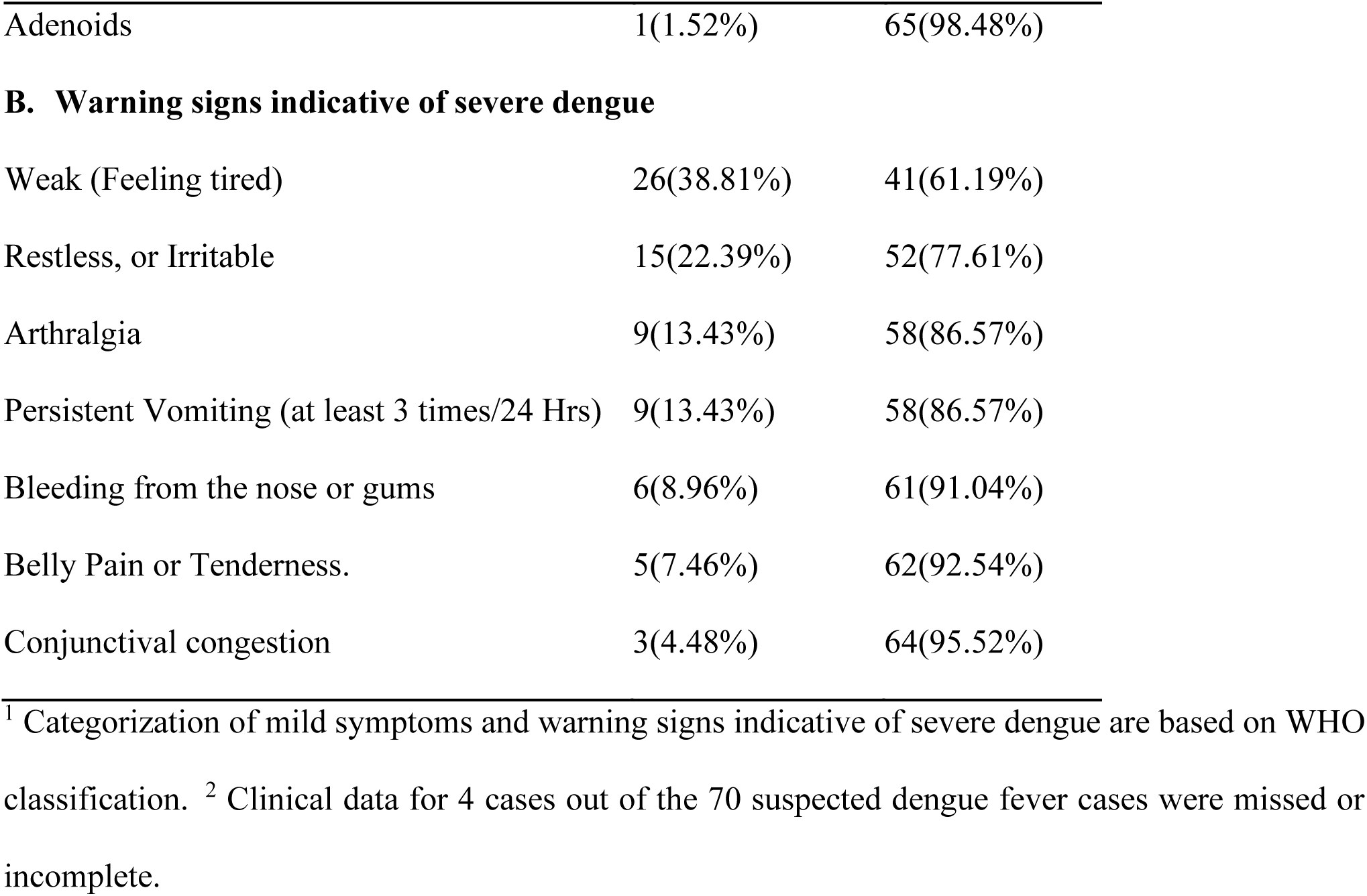
Clinical characteristics of dengue fever suspected cases (n = 70) reported at health facilities.

| Clinical symptoms<sup>1</sup> | Yes<sup>2</sup> | No |
| --- | --- | --- |
| **A. Mild symptoms** |  |  |
| Fever: sudden onset of high fever | 66(98.15%) | 1(1.49%) |
| Chills | 47(70.17%) | 20(29.85%) |
| Vomiting or Nausea | 31(46.27%) | 36(53.73%) |
| Aches and Pains: Typically, behind the eyes, muscle, joint, or bone pain | 31(46.27%) | 36(53.73%) |
| Myalgia | 28(45.9%) | 33(54.1%) |
| Running nose | 21(31.34%) | 46(68.66%) |
| Diarrhoea | 13(19.4%) | 54(89.60%) |
| Rash | 8(13.11%) | 53(86.89%) |
| Sore throat | 6(8.89%) | 61(91.04%) |
| Dizzy | 5(7.46%) | 62(92.54%) |
| Shivering | 4(5.97%) | 63(94.03%) |
| Pruritus | 2(2.99%) | 65(97.01%) |
| Adenoids | 1(1.52%) | 65(98.48%) |
| B. Warning signs indicative of severe dengue |  |  |
| Weak (Feeling tired) | 26(38.81%) | 41(61.19%) |
| Restless, or Irritable | 15(22.39%) | 52(77.61%) |
| Arthralgia | 9(13.43%) | 58(86.57%) |
| Persistent Vomiting (at least 3 times/24 Hrs) | 9(13.43%) | 58(86.57%) |
| Bleeding from the nose or gums | 6(8.96%) | 61(91.04%) |
| Belly Pain or Tenderness. | 5(7.46%) | 62(92.54%) |
| Conjunctival congestion | 3(4.48%) | 64(95.52%) |

<sup>1</sup> Categorization of mild symptoms and warning signs indicative of severe dengue are based on WHO classification. <sup>2</sup> Clinical data for 4 cases out of the 70 suspected dengue fever cases were missed or incomplete.

### 2.3. Blood sample collection and processing

Each of the 70 dengue-suspected patients submitted blood samples for laboratory diagnosis of the causative agent for febrile illness (Supplementary Table 1). The blood samples were allowed to clot, and the sera were separated. The sera were then shipped in a triple package cold chain to the National Arbovirus Laboratory of the EPHI. At the laboratory, the sera were kept frozen at −20°C until they were used for DENV viremia detection using reverse transcriptase (RT)-PCR (RT-PCR).

### 2.4 Viral RNA Extraction and PCR Detection of DENV

Total RNA was isolated from each serum sample using the QIAmp viral RNA kit (QIAGEN, INC, Valencia, CA). The RNA was eluted in 50 μl of AVE buffer, which was provided with the kit. DENV Viremia Detection was done using RealStar® Dengue RT-PCR kit (Altona Diagnostics, Hamburg-Altona, Germany). Briefly, 20 µl of the master mix (containing primers and probes) was mixed with 10 µl of the RNA sample or control. The mixture was centrifuged for 30 seconds at approximately 1000 x g (∼3000 rpm). The reactions were incubated for 20 minutes at 55 °C, followed by 40 cycles of 95°C for 1 min, 55°C for 45 seconds, 72°C for 15 seconds, and final incubation at 68°C for 10 min. The PCR results were interpreted based on the manufacturer’s protocol. A threshold cycle (Ct) value of ≤ 36 for DENV-specific RNA and a Ct value of ≤ 30 for the Internal Control were considered positive samples for DENV.

### 2.5 Semi-Nested RT-PCR Amplification (Amplicon size 511 bp)

RNA samples with a Ct-value of 31 or lower and sufficient volume were selected for CprM RT-PCR amplification following the procedure described elsewhere [12]. Briefly, the DENV CprM region (652) bp was first amplified using forward primer (FP): 5′-TCAATATGCTGAAACGCGCGAGAAACCG-3′ (132– 153) and the reverse primer (RP): 5′-GCGCCTTCNGNNGACATCCA -3′ (764–783) and the high-capacity cDNA reverse transcription kit (Invitrogen). The reactions were incubated first at 98°C for 10 seconds followed by 35 cycles of 98°C for 10 seconds, 60°C for 20 seconds, 72°C for 30 seconds, and final incubation at 72 °C for 5 minutes. Semi–nested PCR was also used to amplify 511 bp of the CprM region of the viral genome using amplified product as a template, and the same FP: 5′-TCAATATGCTGAAACGCGCGAGAAACCG-3′ (132–153) but a different RP-: 5’-TTGCACCAACAGTCAATGTCTTCAGGTTC-3’ (614-642)[12]. The semi-nested amplification was performed in an Applied Biosystems Veriti 96-Well Thermal Cycler (ThermoFisher Scientific) using the following cycling parameters: 94°C for 2 min; 40 cycles of 94°C for 30 s, 70°C for 1 s (ramp rate 20%), 55°C for 45 s (ramp rate 20%), and 65°C for 3 min, 20 s; and final extension for 10 min at 65°C [13]. Following the amplification, 5 μl of the first amplicon and semi-nested amplicon size of 652 bp and 511 bp, respectively were visualized on ethidium bromide-stained agarose gel under UV light. Only the 511 bp DENV amplified PCR products were purified using a QIAquick PCR purification Kit (QIAgen, Germany) and were sequenced in both strands using a Big Dye Terminator Cycle Sequencing kit (Applied Biosystems), and the same primers used for nested-PCR. The CprM sequences were confirmed by BLAST. The forward and reverse sequences were aligned and manually edited using Bio Edit software to obtain the consensus sequence. The obtained sequences were analyzed using Dengue Virus Typing Tool at Genome Detective server [13] to identify the serotypes, genotypes, and lineages of DENV associated with the outbreak.

### 2.6 Phylogenetic analysis

The CprM consensus sequences for 21 DENV strains from Dire Dawa City were deposited in GenBank under accession numbers PP751832-PP751852. Nucleotide BLAST searches were performed for each of the 21 new sequences, and all DENV-3III CprM gene and whole genome sequences with >98% similarity were downloaded. Duplicates and identical sequences from the same country and collection date were removed. Representative sequences for DENV-3 genotypes I, II, and V were also included as references. The final data set include a total of 123 DENV-3 CprM gene sequences collected between 2006-2023 across the globe.

For phylogenetic reconstruction, sequences were aligned using MAFFT (version 7.520)[14], and a maximum likelihood tree was constructed with the IQ-TREE software (version 2.2.2.7)[15] using the TIM2e+G4 substitution model (according to AICc) with a bootstrap value of 10,000. Finally, the resulting tree was visualized and annotated using iTOL (Interactive Tree Of Life)[16].

## 3. Results

### 3.1. Patient characteristics

Among the 70 suspected dengue fever (DF) patients, 38 (54%) were female, as indicated in Table 1 and Table S1. The median age was 30 with an interquartile (IQ) range of 17 years. Most (81%) of the suspected cases visited health facilities as outpatients (Table 1 and Supplementary Table 1), while the remaining 13 cases required inpatient hospitalization for a duration of 2 to 10 days (Supplementary Table 1).

**Table 1.** Status of admission and demographic characteristics of dengue fever suspected cases (n = 70) reported at health facilities in the Dire Dawa City Administration, 2023.

| Variable | Specific variable | N (%) |
| --- | --- | --- |
| Age | Median age 30; IQR 17 | - |
| Sex | Male | 32(46) |
|  | Female | 38(54) |
| Patient admission status | Outpatient | 57(80) |
|  | Inpatient | 13 (20) |

The clinical signs/symptoms observed in these (n = 70) DF suspected cases, whether admitted as outpatients or inpatients, covered a wide range of symptoms consistent with febrile illness. Nearly all of them exhibited three or more clinical symptoms consistent with mild DF (Supplementary Table 1). However, regardless of hospitalization status, 32 (45.7%) of these patients displayed one or more early warning signs indicative of severe dengue[11]. Among the 13 patients who were hospitalized for 2 to 10 days, all except two had at least one symptom indicative of severe dengue (Supplementary Table 1). The most commonly observed clinical presentations were acute fever (n = 66; 98.15%)), headache (n = 65; 97.01%)) and chills (n = 47; 70.17%)) followed by aches and pains (n = 31; 46.27%) and vomiting/nausea (n =31; 46.27%) (Table 2). In addition, all of the cases with early warning signs of severe dengue were later confirmed to harbor DENV viremia by RT-PCR. Supplementary Table 1 summarizes the details of the demographic and clinical characteristics of study participants.

### 3.2. DENV Viremia Detection

Among 70 specimens, 3 samples failed to pass the DENV nucleic acid amplification quality control. Among the 67 samples that passed quality control, 65.67% (44/67) had DENV RNA detected(Figure 2).

Supplementary Table 2 summarizes laboratory investigation of dengue-confirmed patients, including DENV serotypes, genotypes, linage, and sublineages with their corresponding GenBank Accession Numbers.

### 3.3. DENV sequence classifications

Based on rRT-PCR CT <31, 27 samples out of 44 DENV viremia-positive samples were selected for CprM sequencing, and 21 were successful (Figure 2) allowing classification of their serotypes, genotypes, and lineages (Supplementary Table 2). The sequence quality for 6 samples was not sufficient, and the DENV serotypes could not be determined (Figure 2). According to the new proposed DENV nomenclature [13], the Dengue Virus Typing Tool assigned all 21 sequences to DENV-3, genotype III, major lineage B (DENV-3III_B), and the minor lineages were assigned as follows: 17 sequences belonged to or were most closely related to minor lineage 2 (DENV-3III_B.2), and 4 sequences belonged to or were most closely related to minor lineage 3 (DENV-3III_B.3) (Figure 3).

**Figure 3.**
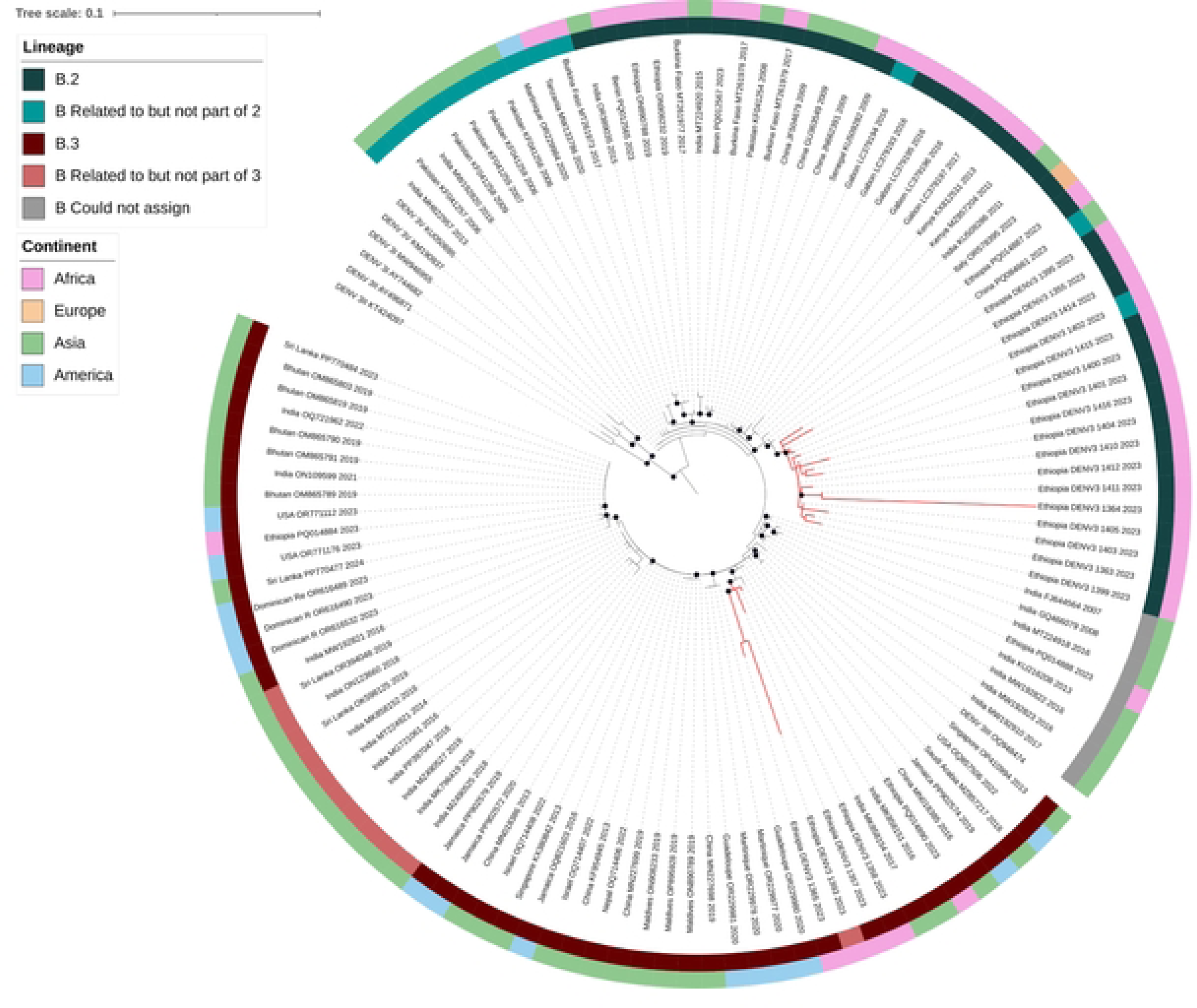
Maximum likelihood phylogenetic tree of DENV-3 genotype III (DENV-3III) strains isolated during the 2023 outbreak in Dire Dawa, Ethiopia. The tree includes CprM sequences newly generated in this study (red branches, N = 21) and representative reference sequences from DENV-3III (black branches, N = 102). Sequences are labeled with the accession number, country of origin, and year of isolation, and the outer circles indicate continent of origin and DENV-3III_B minor lineage. Nodes with ultrafast bootstrap values of 95 and above are indicated with black circles. The internal tree scale represents the nucleotide substitutions per site.

### 3.4 Phylogenetic analysis

We investigated phylogenetic relationships between the 21 new DENV-3III_B CprM sequences obtained in this study and other publicly available DENV-3III_B CprM sequences from various regions including Africa (Kenya, Gabon, Burkina Faso, Mozambique, Tanzania, and Ethiopia), Asia (Middle East, India, Sri Lanka, Thailand, and China), the Americas (Jamaica, Martinique, Dominican Republic and USA), and Europe (Italy).

As expected, the 17 DENV-3III_B minor lineage 2 (DENV-3III_B.2) sequences clustered separately from the 4 DENV-3III_B minor lineage 3 (DENV-3III_B.3) or related sequences (Figure 3). The DENV-3III_B.2 sequences from our study clustered with other sequences from 2023 collected across different continents, including one from an outbreak in Lazio, Italy, and one from China attributed to a case imported from Burkina Faso. On the other hand, the DENV-3III_B.3 sequences clustered with two sequences from India obtained in 2016 and 2017. Importantly, both the DENV-3III_B.2 and DENV-3III_B.3 clusters contained a reference sequence isolated from the 2023 outbreak in the Afar region of Ethiopia[17], supporting the hypothesis that the outbreak in Dire Dawa was related to the outbreak in the Afar region, and indicating that multiple lineages co-circulated in both outbreaks.

## 4. Discussion

Knowledge of the genetic diversity of DENV strains responsible for outbreaks is essential for efficient management, vaccine development, and outbreak prevention. Though Ethiopia’s eastern region, including Dire Dawa City, has seen many DF outbreaks since 2013, there is scanty information regarding the genetic diversity of circulating DENV strains. In this study, we used genetic analysis of the DENV CprM region to demonstrate that the 2023 outbreak in Dire Dawa City was due to two co-circulating lineages of DENV-3III_B, each of which was closely related to sequences from a related outbreak in the Afar region of Ethiopia in 2023. Our results are consistent with a recent study that analyzed 7 full DENV-3III genome sequences from the Afar outbreak and found evidence for multiple introductions. However, the paucity of regional DENV sequence data makes it difficult to determine the timing and origin of these introductions. Recently, other DENV-3III outbreaks have been reported in East Africa, notably in South Sudan in 2022[18] and in Kenya in 2019[19]. We were not able to assess the genetic relatedness between the current strains and outbreak strains from these neighboring countries due to the fact they employed other targets for sequencing such as the E gene. These results underscore the need for further genomic surveillance of DENV in East Africa, ideally using full-length genome sequences.

In contrast to the prior dengue outbreaks in Ethiopia which were more mild[7],[20],[21],[22], in this study 46% (n = 32) of the 70 DF-suspected cases displayed early warning signs indicative of severe dengue, while the remaining cases showed mild symptoms. All of these cases with early warning signs of severe dengue were confirmed to harbor DENV viremia by RT-PCR.

We propose three scenarios that might have led to the occurrence of severe cases of DF during the 2023 outbreak in Dire Dawa City. One possibility is the observed severe dengue cases could be secondary DENV-3III infections that happened after primary infection with a different serotype. In support of this, prior DF outbreaks reported from Dire Dawa City were associated with DENV-1 and DEV-2 strains [7],[20],[21],[22],(Mesfin et al, in Press Viruses, MDPI), although previous attempts to determine serotypes were restricted to a small number of DENV strains. Thus, the recurrent occurrence of dengue outbreaks associated with different DENV serotypes calls for the need for close monitoring and management of future DF outbreaks to effectively control and manage secondary infections.

The second possibility is that DENV-3III is more pathogenic, as suggested by prior research that connected outbreaks of DENV-3III to Dengue Hemorrhagic Fever (DHF) and Dengue Shock Syndrome (DSS) [23].

The third possibility is that the severe cases may be the result of co-infection with malaria parasites, as both infections are co-endemic in Dire Dawa City, and there is a chance that co-infection can worsen disease severity and outcomes. This is supported by the recent surge in malaria[24],[25] and dengue cases[26],[27] in Dire Dawa City which posed significant challenges for healthcare providers. Given the co-endemicity and overlapping of symptoms (e.g., fever), there is a risk of co-infection with both dengue and malaria and misdiagnosing dengue as malaria (and vice versa). Thus, an integrated approach involving surveillance, education, and differential diagnosis is crucial for better patient care in Dire Dawa City Administration. Healthcare professionals need to be aware of the clinical features, diagnostic tests, and management protocols for malaria and dengue. In addition, the Dire Dawa City Administration should avail appropriate laboratory tests (such as rapid diagnostic tests and PCR) at all health facilities that facilitate accurate diagnosis of dengue and malaria and prompt treatment and management of severe cases.

Among the five distinct genotypes (I–V) of DENV-3, all 21 sequenced DENV strains from the present study belonged to DENV-3III, which is the most widespread and was associated with large outbreaks in Asia, Africa, and the Americas[28],[29],[30]. The first autochthonous case of DENV-3III in Africa was reported in Mozambique in 1985[31]. Several man-made and natural factors, including (i) migration and conflict-related population displacement, (ii) frequent migration via trade routes that can spread the virus to new areas, and (iii) recurrent flooding that is made worse by climate change that provides ideal breeding grounds for Aedes mosquitoes may have contributed to the rapid geographic expansion of DENV-3III in Africa, resulting in outbreaks in areas like Dire Dawa City[3],[28],[32],[23],[33].

Even though this work offers important insights into DENV strains responsible for the 2023 DF outbreak in Dire Dawa City where severe cases and deaths were recorded, it suffers limitations. First, the study’s capacity to detect genetic variation was limited due to the use of a short genomic target (CprM, 511 base pairs) for the characterization of the outbreak strains. Expanding the genomic coverage could have enhanced our understanding of DENV diversity and relationships to other DENV-3III strains across the globe. Second, DENV CprM sequencing was performed on a small proportion of DF-suspected cases reported during the outbreak, and a larger sample size may have identified greater genetic diversity. Third, the study lacks information on other febrile illness-causing pathogens like malaria parasite examination with microscopy or rapid diagnostic test results, for the studied DF-suspected cases, so cases of co-infection may have been missed. Future research addressing these limitations will therefore improve our comprehension of disease dynamics, provide a thorough understanding of DENV strains and their evolutionary history, expand our understanding of DENV outbreaks, and provide guidance for more effective preventative measures.

## 5. Conclusions

In summary, our results demonstrate that the 2023 DF outbreak in Dire Dawa City, Ethiopia, was due to multiple co-circulating lineages of DENV-3III. The severe cases recorded during this outbreak may be due to either the genetic makeup of DENV-3III, secondary DENV infection, or co-infection of DENV and malaria. Additionally, the intercontinental transmission of DENV-3III underscores the importance of vigilant surveillance and preparedness to mitigate the impact of such outbreaks. Thus, collaborative efforts across borders are crucial for effective prevention and control strategies.

## Supplementary Materials

Supplementary Table 1: Demographic and clinical parameters of dengue suspected patients and Supplementary Table 2: Laboratory investigation of dengue confirmed patients, including DENV serotype and genotype with their corresponding GenBank Accession Numbers.

## Author Contributions

Conceptualization, A.A.N. and S.A.A.; study design, S.A.A., A.Mu. And T.G.; methodology, A. Mu., S.A.A., T.G., H.A.; software, T.G.; validation, T.G., DM.RG., A.A.N., and Z.Z.; formal analysis, T.G., DM.RG., and A.Ay.; Laboratory investigations, D.H.A., T.K.W., G.B., A.H.B, D.T., E.A, EZA, A.As, A.K.K., K.M., and R.J.; resources, A.Mu, A.Mi., S.A.A., A.Ab., G.T., M.H; Field data and samples acquisition, E.Z.A., D.T., and A.K.K. data curation, T.G., and A.A.N.; writing—original draft preparation, T.G., and A.A.N.; writing—review and editing, T.G., A.P., A.Mu., and A.A.N.; visualization, T.G., DM.RG, A.Ay., and A.P.; supervision, S.A.A. All authors have read and agreed to the published version of the manuscript.

## Acknowledgments

We would like to thank all study participants in this study. We are also grateful for the support and cooperation we received from physicians, nurses of Dire Dawa City health facilities, and administrative authorities of Dire Dawa City Administration Health Hureau and the health facilities administration. The authors acknowledge EPHI’s support of the field study and AHRI and EPHI’s reagent support of the molecular analysis.

## Data Availability Statement

The datasets generated and/or analyzed during the current study are available in the manuscript and the Supplementary Materials. The sequence data presented in this study are openly available in the GenBank database, accession numbers PP751832-PP751852.

## Disclaimer/Publisher’s Note

The statements, opinions, and data contained in all publications are solely those of the individual author(s) and contributor(s) and not of the journal and/or the editor(s) and should not be construed as official, or as reflecting true views of the Armauer Hansen Research Institute and Ethiopian Public Health Institute.

